# Laboratory changes associated with medication non-adherence in patients with hypertension over six months of the COVID-19 pandemic

**DOI:** 10.1101/2022.03.15.22272192

**Authors:** Shu-Mei Chang, Yi-Chun Chen, Chin-Feng Hsuan, I-Cheng Lu, Hung-Yi Chuang

## Abstract

The coexistence of multiple diseases is common in the elderly and often accompanied by medication non-adherence. This study investigated the relationship between medication non-adherence and laboratory findings inpatients with hypertension and hypertensive comorbidities (i.e., diabetes and nephropathy) in southern Taiwan during 6 months of the coronavirus disease pandemic. This was a panel study and involved outpatients from three hospitals classified as regional hospitals or above. Questionnaireswere usedto collect information on patient demographics, diet, medication adherence, and laboratory data at the time of recruitment and 6 months after. A total of 140 patients with only hypertension and 98 patients with hypertension and comorbidities were recruited, and the changes inblood pressure andlaboratory data were assessed after 6 months. Analyses performed with generalized estimating equations showed that patients who had not forgotten to take medication had a higher estimated glomerular filtration rate. Moreover, patients who did not change their medication time arbitrarily had lower low-density lipoprotein levels. Furthermore, patients who did not stop or interrupt their medication arbitrarily had lower diastolic blood pressuresand low-density lipoprotein levels. Overall, patients with better medication adherence had better estimated glomerular filtration rates,lower low-density lipoprotein levels, and lower diastolic blood pressures.

## Introduction

The present study was conducted in 2020, during the coronavirus disease (COVID-19) pandemic, when most peopleavoided going to public places and even visits to hospitals were reduced. A study in Malaysia showed that patients with hypertension were more likely to be negatively affected by the pandemic [1]. According to the results of the National Nutrition and Health Survey conducted from 2017–2020, approximately 5.23 million people above the age of 20 had hypertension, accounting for about a quarter of the population of Taiwan [2].

Hypertension is the main risk factor for many global illnessesand premature death, and its burden of disease has exceeded that of smoking and overweight [3]. Hypertension is also a common risk factor for major chronic diseases such as cardiovascular disease, stroke, diabetes, and nephropathy. In 2010, high blood pressure was the leading cause of death and disability-adjusted life years worldwide [4]. Moreover, in 2017, the American Heart Association reduced blood pressure threshold values for hypertensiontopromote earlier control of comorbidities andreduce the incidence of stroke and cardiovascular disease [4]. Hypertension is an important global health challenge and a leading preventable risk factor for premature death and disability worldwide [5].

Most patients with hypertension require more than two different kinds of anti- hypertensive drugs to maintain their blood pressure within the ideal range (below 140/90mmHg) as well as to reduce the incidence of comorbidities, such as cardiovascular disease, cerebrovascular disease, diabetes, chronic kidney disease, and retinopathy [6,7].

In current clinical hypertension management practices, the main obstacles in managing patients with hypertension are comorbidity management and poor medication adherence, which may be explainedby a lack of persistence[8]. The World Health Organization has recently highlighted increased adherence as a key developmental need in reducing cardiovascular disease [5], and researchingnew, effective medications is required to better control hypertension in patients. However, in the absence of new, effective anti-hypertensive medications, improving medication adherence of readily available therapies is important and has been emphasized by recent guidelines [9]. Approximately half of patients with uncontrolled hypertension do not take their medication correctly [10]. In the 2015 US National Health Interview Survey,23.8 million adults with hypertension reported an overall weighted non-adherence rate of 31.0% [11]. Similarly, an analysis of data from Taiwan’s National Health Insurance Research Database in 2010 revealed that up to 85.5% of patients with poor disease control are non-adherent to treatment[12].

Furthermore, according to Schiffrin et al.[13], the most common comorbidities in patients with coronavirus disease (COVID-19) and acute respiratory distress syndrome were hypertension (27%), diabetes (19%), and cardiovascular disease (6%).Although there have not yet been severe COVID-19 outbreaks in Taiwan, patients with hypertension and hypertensive comorbidities are still inevitably involved in behaviors that include medication non-adherence. Therefore, the purpose of this study was to investigate the changes in laboratory data and medication non-adherence among patients with hypertension and hypertensive comorbidities (i.e., diabetes and nephropathy). By doing this, we hoped to understand whether the pandemic impacted patients’ willingness to visit doctors and take medications. The findings may serve as a guide for betterpatientadherence as well as treatment adjustmentsand enhanced patient education by clinical physicians.

## Materials and Methods

### Patient Recruitment

A panel study was conducted between January 2020 and June 2021. During the 6-month study period,the COVID-19 pandemic was ongoingand patient visits to hospitals had reduced. We recruited outpatients with hypertension and hypertensive comorbidities, such as diabetes or nephropathy, in three hospitals classified as regional hospitals or above. The patients were recruited into two groups:those with hypertension requiring medication (n=140)andthose with hypertension and other comorbidities (i.e., diabetes or nephropathy) (n=98).

The study plan was reviewed and approved by the Research Ethics Committee (IRB: EMRP108065). This study complied with research ethics regulations and the principle of informed consent. The consent form was presented to the subjects in a quiet room, the contents of the consent form were explained, and questions regarding the study were answered in a way that the subjects could understand. We confirmed that the subjects understood the contents of the research and gave them sufficient time to consider and communicate. Finally, when the subjects agreed to participate in the study, they signed the consent form.The study was explained to the patients who met the inclusion criteria during their visit to their doctors, after which, those who agreed signed the consent form and completed the questionnaire anonymously in a quiet and empty room.

### Inclusion Criteria

Outpatients who were at least 40 years old and had been diagnosed with hypertension by a physician at least 1 year prior were included in the study. Moreover, the ability to take drugs alone; clear consciousness;the ability to communicate with the interviewer in the Mandarin, Taiwanese, or Hakka languages; and the ability to understand verbal questions were also inclusion criteria. Patients with a terminal-stage disease with a life-expectancy of 6 months or less, or other special circumstances, such as aggressiveness, were excluded.

### Questionnaires and Interviews

The questionnaire was specifically tailored to meet the purpose of this study and was based on references from both domestic and foreign literature [14]. Several revisions were made according to expert discussions, and the draft was finalized after a preliminary trial. The questionnaire had seven parts: (1)basic information, including sex, age, religion, marital status, living conditions, body weight, and height; (2)medication status, includingthe number of pills taken, medication frequency, regularity in taking medications, and presence/absence of side effects;(3)medication non-adherence behavior was recorded, includingforgetting to take medication more than two times in a week within the past 3 months, after which they were divided into groupsthat includedforgetting to take medication, changing the medication time arbitrarily, stopping or interrupting the medication arbitrarily, reducing the medication times arbitrarily, decreasing the dosage arbitrarily, and purchasing medications not prescribed by physicians arbitrarily [14]; (4)follow-up visits, including whether or not the patient came for follow-ups regularly;(5) lifestyle, such as drinking, smoking, coffee consumption, exercise, sleep, dietary supplements, and tea consumptionhabits;(6) past medical history and self-assessed health status; and (7)diet, including types of diet[15].

### Blood pressure measurement and laboratory investigations

Blood pressures (systolic and diastolic values), glycated hemoglobin (HbA1c), fasting blood glucose (FBS), creatinine, the urine protein/urine creatinine ratio, estimated glomerular filtration rate (eGFR), and low-density lipoprotein levels (LDL) were measured initially and during patient follow-ups 3 to 6 months after, upon orders from the clinical physician. Laboratory investigations were performed in the same laboratory to avoid bias.

### Data Analysis

Descriptive statistics was used to analyze the distribution of demographic variable data. Categorical variables were presented in frequency tables and expressed as percentages. Continuous variables were expressed as average values with standard deviations. The chi-square (χ^2^) and Fisher’s exact tests were used to compare two categorical variable groups. Two-tailed independent sample t-tests and paired sample t-testswere used to compare two continuous variable groups. Three continuous variable groups were compared using a one-way analysis of variance (ANOVA). This study used generalized estimating equations (GEEs) to find the association between medication non-adherence behaviors and laboratory data adjusted interference factors (sex, age, education, and marital status), and data from the same participants were analyzed multiple times. SPSS18.0 statistical software was used for data analysis, and α was set at0.05.

## Results

A total of 238 patients were recruited: 140 with only hypertension, 59 with hypertension and diabetes, and 39 with hypertension and nephropathy. There were no significant differences in age, body mass index, sex, marital status, occupation, and living condition between the three groups. Regarding education, there were 35 (25.0%) only hypertension patientsthat had college degrees, 15 (25.4%) hypertension and diabetes patients with college degrees, and 18 (47.4%) hypertension and nephropathy patients with college degrees. The proportion of people with college degrees with hypertension and nephropathy was significantly higher than the proportion of people with college degrees in the other two groups(P=0.016) (Table 1).

**Table 1.** Demography of patients with only hypertension, with hypertension and diabetes, and with hypertension and nephropathy (n=238)

| **Variables** | **Only**<br>**Hypertension**<br>**(%)** | **Hypertension**<br>**and Diabetes**<br>**(%)** | **Hypertension**<br>**and**<br>**Nephropathy**<br>**(%)** | **n** | **p-**<br>**value** |
| --- | --- | --- | --- | --- | --- |
| **Age, years**<br>M(SD)<sup>b</sup> | 61.8(11.1) | 64.2(8.9) | 60.9(12.7) | 238 | 0.376 |
| **BMI<sup>a</sup>, kg/m<sup>2</sup>**<br>M(SD)<sup>b</sup> | 26.7 (4.15) | 27.5(4.36) | 26.4(4.63) | 238 | 0.262 |
| **Sex** |  |  |  |  |  |
| Male | 77 (55.0%) | 32 (54.2%) | 21 (53.8%) | 130 | 0.996 |
| Female | 63 (45.0%) | 27 (45.8%) | 18 (46.2%) | 108 |  |
| **Marital status** |  |  |  |  |  |
| Married | 103 (74.1%) | 45 (76.30%) | 27 (69.2%) | 174 | 0.816 |
| Single | 27 (19.4%) | 10 (16.9%) | 11 (28.2%) | 48 |  |
| Divorced | 6 (4.3%) | 3 (5.1%) | 1 (2.6%) | 10 |  |
| Widowed | 3 (2.2%) | 1 (1.7%) | 0 (0.0%) | 4 |  |
| **Religion** |  |  |  |  |  |
| Taoism | 82 (61.7%) | 26 (44.8%) | 17 (48.6%) | 125 | 0.062 |
| Buddhism | 36 (27.1%) | 27 (46.6%) | 9 (25.7%) | 72 |  |
| Christian | 7 (5.3%) | 3 (5.2%) | 4 (11.4%) | 14 |  |
| Catholic | 1 (0.8%) | 0 (0.0%) | 0 (0.0%) | 1 |  |
| Others | 7 (5.3%) | 2 (3.4%) | 5 (14.3%) | 14 |  |
| **Education** |  |  |  |  |  |
| Junior<br>Highschool | 50 (35.7%) | 29 (49.2%) | 8 (21.1%) | 87 | 0.016 |
| Senior/Vocational<br>highschool | 44 (31.4%) | 12 (20.3%) | 12 (31.6%) | 68 |  |
| College | 35 (25.0%) | 15 (25.4%) | 18 (47.4%) | 68 |  |
| Master | 11 (7.9%) | 3 (5.1%) | 0 (0.0%) | 14 |  |
| **Occupation** |  |  |  |  |  |
| Employed | 49 (35.0%) | 20 (33.9%) | 12 (30.8%) | 81 | 0.885 |
| Unemployed | 91 (65.0%) | 39 (66.1%) | 27 (69.2%) | 157 |  |
| **Living** |  |  |  |  |  |

| Condition |  |  |  |  |  |
| --- | --- | --- | --- | --- | --- |
| Lives Alone | 10 (7.1%) | 4 (6.9%) | 5 (12.8%) | 19 | 0.154 |
| Lives with Family | 130 (92.9%) | 54 (93.1%) | 33 (84.6%) | 217 |  |

<sup>a</sup>Body mass index
<sup>b</sup>Mean (standard deviation)

Patients were divided into different medication non-adherence behavior groups. Among the patients who did not return after 6 months, 4 (2.9%) had only hypertension, 3 (5.1%) had hypertension and diabetes, and none (0.0%) had hypertension and nephropathy. The difference among the groups was not significant (P=0.344)(Table 2).

**Table 2.** Medication non-adherence behavior and loss to follow-up among patients with only hypertension, with hypertension and diabetes, and with hypertension and nephropathy (n=238)

|  | Only Hypertension | Hypertension and Diabetes | Hypertension and Nephropathy | n | p-value |
| --- | --- | --- | --- | --- | --- |
| **Forgot to take medication** |  |  |  |  |  |
| **Yes** | 46 (32.9%) | 12 (20.3%) | 10 (25.6%) | 68 | 0.184 |
| **No** | 94 (67.1%) | 47 (79.7%) | 29 (74.4%) | 170 |  |
| **Changed medication time arbitrarily** |  |  |  |  |  |
| **Yes** | 24 (17.1%) | 5 (8.5%) | 7 (18.4%) | 36 | 0.248 |
| **No** | 116 (82.9%) | 54 (91.5%) | 31 (81.6%) | 201 |  |
| **Stopped or interrupted medication arbitrarily** |  |  |  |  |  |
| **Yes** | 16 (11.4%) | 3 (5.1%) | 3 (7.7%) | 22 | 0.346 |
| **No** | 124 (88.6%) | 56 (94.9%) | 36 (92.3%) | 216 |  |
| **Reduced the medication times arbitrarily** |  |  |  |  |  |
| **Yes** | 9 (6.4%) | 1 (1.7%) | 2 (5.3%) | 12 | 0.379 |
| **No** | 131 (93.6%) | 58 (98.3%) | 36 (94.7%) | 225 |  |
| **Decreased the dosage arbitrarily** |  |  |  |  |  |
| **Yes** | 8 (5.8%) | 9 (15.3%) | 4 (10.3%) | 21 | 0.097 |
| **No** | 130 (94.2%) | 50 (84.7%) | 35 (89.7%) | 215 |  |
| **Returned after six months** |  |  |  |  |  |
| **No** | 4 (2.9%) | 3 (5.1%) | 0 (0.0%) | 7 | 0.344 |
| **Yes** | 136 (97.1%) | 56 (94.9%) | 39 (100.0%) | 231 |  |

The patients were followed for 6months, and blood laboratory data were analyzed using a one-way ANOVA. The average HbA1c of patients with hypertension and diabetes was 6.9 (1.65), which was significantly higher than that of the other two groups (P=0.006). The average creatinine level in patients with hypertension and nephropathy was 2.4 (3.27), which was significantly higher than that of the other two groups (P<0.000). The estimated glomerular filtration rate (eGFR) of patients with hypertension and nephropathy was 59.7 (34.6), which was significantly lower than that of the other two groups(P<0.001) (Table 3).

**Table 3.**
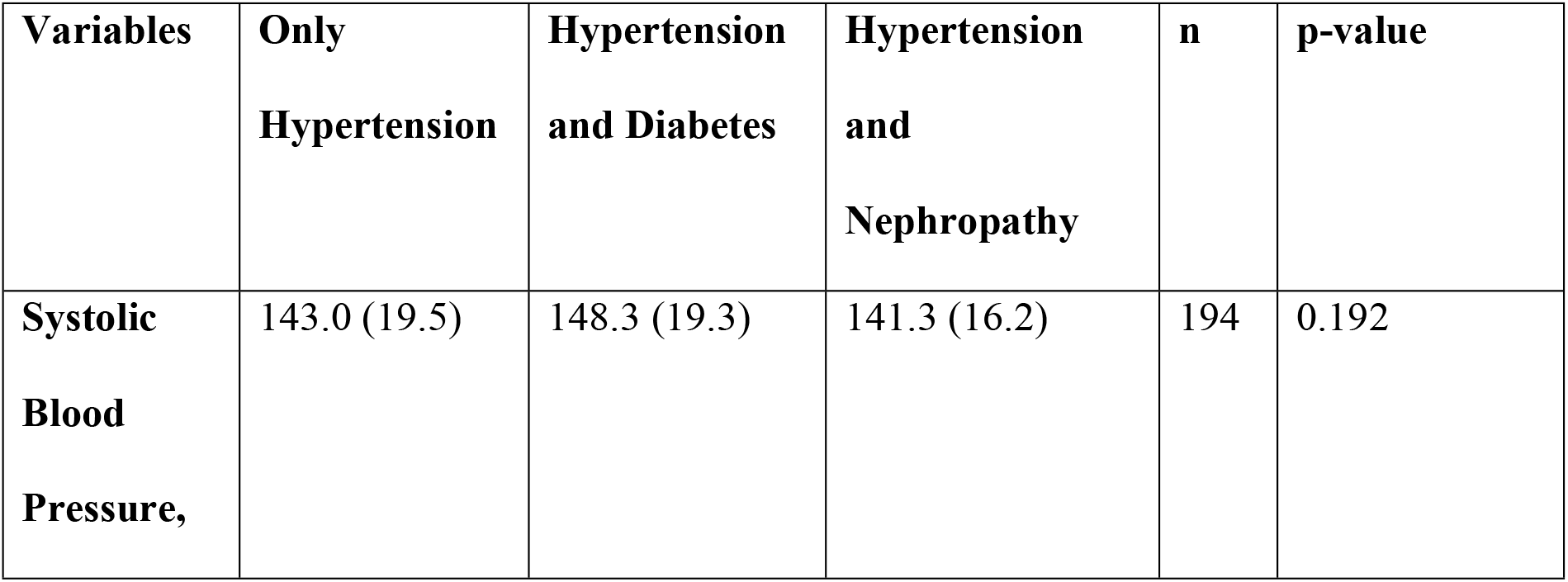
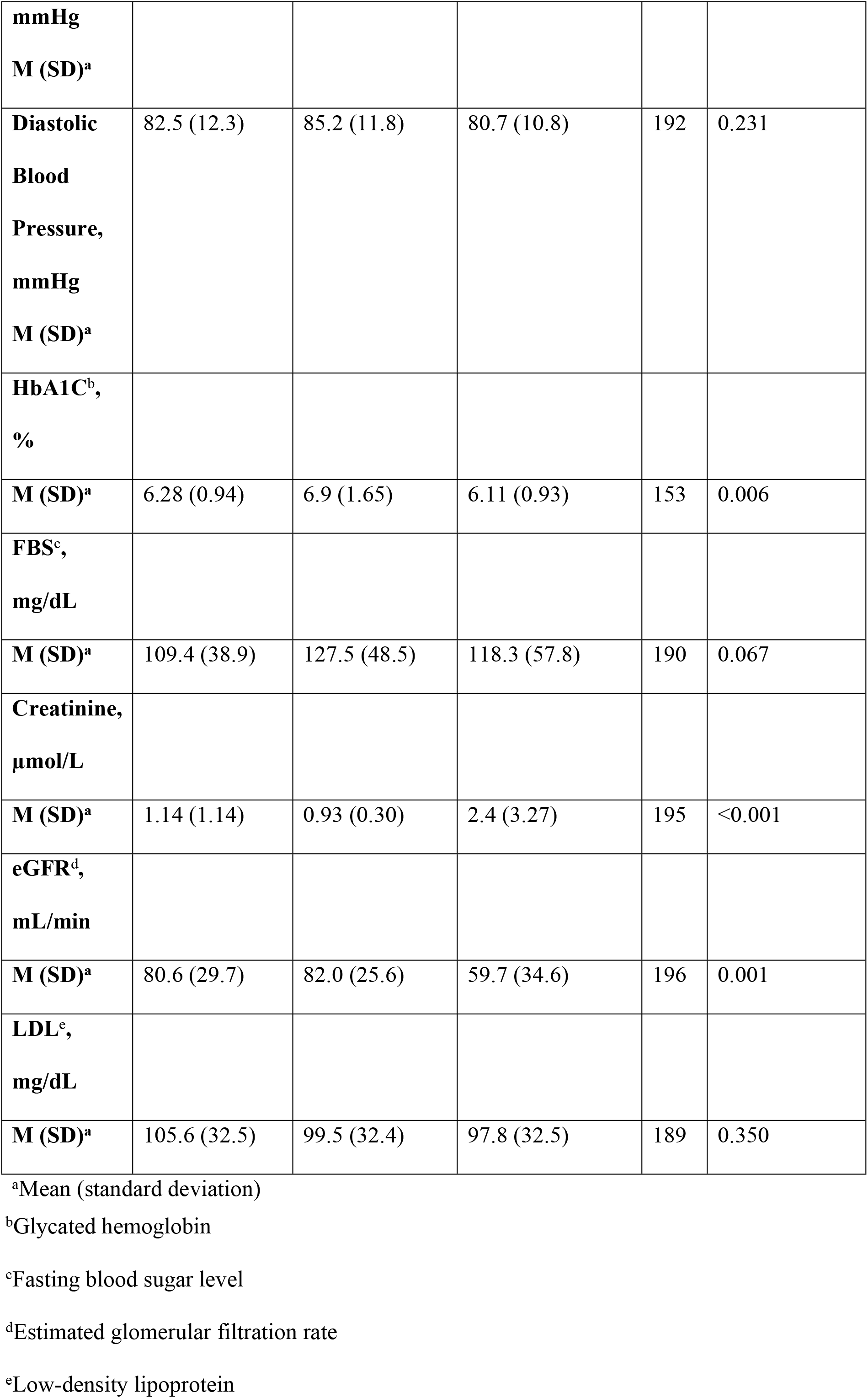
Laboratory data of patients with only hypertension, with hypertension and diabetes, and with hypertension and nephropathy.

After monitoring the changes in blood pressure and laboratory data for 6 months, GEEs were applied according to the non-adherence items collected at the time of the questionnaire, and factors including sex, age, education, marital status were controlled. The eGFR and LDL could be evaluated using parameters of whether or not the patient had forgotten to take medication. The estimated β value was9.887, and the P value was0.024, indicating that eGFR increased in patients who had not forgotten to take medication. With the estimated β value of-13.34 and P value of0.014, LDL levels decreased in patients who did not change their medication time arbitrarily. Moreover, diastolic blood pressures and LDL levels could be evaluated using parameters of whether the patients had stopped or interrupted their medication. The estimated β value was-7.046, and the P value was 0.005, indicating that patients who did not stop or interrupt their medication arbitrarily had lower diastolic blood pressures. Similarly, with an estimated β value of-19.80 and P value of 0.006, patients who did not stop or interrupt their medication arbitrarily had lowerLDL levels.

## Discussion

In the three groups of patients (only hypertension, with hypertension and diabetes, and with hypertension and nephropathy), the proportion of college degrees in the group with hypertension and nephropathy was significantly higher than that of the other two groups. The overall rate of loss to follow-up after 6 months in the three groups was 2.94%. However, the group with hypertension and nephropathy had a return rate of 100%, which may be related to the higher education level of this group. Among older patients with hypertension who have a low level of education and no social support, there is a need for tailored educationto better understand and adhere to medication[16]. Therefore, health education should be increased for patients with only hypertension and patients with hypertension and diabetes to facilitate the control of chronic diseases.

Among patients who changed their medication time and who stopped or interrupted their medication arbitrarily, the group with only hypertension had higher rates of medication non-adherence behavior, which may indicate a lack of insight to the disease. Therefore, health education about medication adherence for this group should be increased to avoid future comorbidities. However, adherence increases with the number of chronic diseases, while sex, age, and number of drugs do not show a consistent effect [17].

Although this study did not find significant differences in medication non-adherence behavior and return rates after 6 months among the three groups, the proportion of patients who forgot to take medication was highest in the group with only hypertension. The rate of stopping or interrupting one’s medication arbitrarily as well as average LDL levels in this group were also higher than that in the other two groups. Therefore, increased risk of cardiovascular disease should be monitored. The group with hypertension and diabetes had lower rates of medication time change and arbitrary reduction of medication times when compared to the other groups. This group, however, had the highest rate of arbitrary decrease of dosage and the highest average blood pressure among the three groups, indicating that clinical healthcare professionals may have to strengthen the health literacy level and promote adherence to the prescribed dosage in patients with hypertension and diabetes. Emphasis should be placed on accurately measuring and recording blood pressure to effectively control these patients’ blood pressures and blood sugar levels.

According to the GEEs to examine changes after 6 months, if the rate of forgetting medication was reduced, the eGFR of the hypertensive group could be increased by approximately 9.887 mL/min (P=0.024). Moreover, if the rate of changing medication time arbitrarily was reduced, the LDL level of patients with hypertension could be reduced by approximately 13.340 mg/dL (P=0.014). Finally, the analysis showed that if the rate of stopping or interrupting medication arbitrarily was reduced, the LDL level of the group with hypertension could be reduced by up to 19.801 mg/dL (P=0.006), and the diastolic blood pressure could be reduced by approximately 7.046 mmHg (P=0.005).

Medication non-adherence is a major barrier to treatment and a key obstacle in improving patient prognosis. Persistent adherence is a complex phenomenon, and no single intervention can solve this challenge. The ability of patients to optimally follow treatment plans is frequently compromised by more than one barrier, including social andeconomic factors, the health care team and system, disease characteristics, disease therapies, andother patient-related factors[18]. For example, patients with merchant occupations, physical inactivity, and diabetes mellitus co- morbidity were significantly associated with higher rates of medication non-adherence [19]. Solving the problems related to each of these factors is necessary if adherence to therapies is to be improved.

Research has shown that multidisciplinary team-based care can effectively improve the approach to hypertension control by offering patient-centered treatment, which is associated with better adherence [21,22] and a greater influence on the health of patients with hypertension. This may be provided through adequate education on the use of medicine from pharmacists to improve adherence. A literature review showed that predictors of adherence included trust, communication, and empathy; efforts to improve patients’ understanding of medication benefits; and access and trust in their provider and health system. Improving providers’ recognition and understanding of patients’ beliefs, fears, and values, as well as their own biases is also necessary to achieve increased medication adherence and improved population health [23]. In Taiwan, clinical physicians usually adjust medications and educate patients with hypertension and hypertensive comorbidities according to changes in laboratory findings and blood pressure measurements at the patients’ house or hospital.Furthermore, a study in Japan revealed that higher medication regimen complexity had a higher non-adherence [20]. Using double-combination or triple-combination therapies may be helpful in reducing the treatment dosage and improving medication adherence.

This study had some limitations. First, this was a pilot study conducted at outpatient departments during the COVID-19 pandemic, even though the severity and rate of complications were not high. Second, the decrease in patient visits to the outpatient department due to the COVID-19 pandemic made it difficult to follow up with the patients. We intend to continue data collection and analysis to examine more interesting findings.

## Conclusion

This study was conducted during the COVID-19 pandemic. Analyses performed with generalized estimating equations showed that patients who had not forgotten to take medications had a higher eGFR, and patients who did not change their medication time arbitrarily had lower LDL levels. Moreover, patients who did not stop or interrupt their medication arbitrarily had lower diastolic blood pressures and LDL levels. The results of this study may provide more insight on the relationship between medication non-adherence and changes in laboratory data, which may be helpful in adjusting medicines and enhancing patient education by clinical physicians.

## Data Availability

All relevant data are within the manuscript and its Supporting Information files.

## Acknowledgments

Thanks for our part time worker, Liang-Yu Chen, and the nurse, Yin-Jin Lin, and a lot of people who joined in this study.

